# Acceptability of Rectal Artesunate as pre-referral treatment for severe malaria in children under 5 years by health workers and caregivers in the Democratic Republic of the Congo, Nigeria and Uganda

**DOI:** 10.1101/2021.12.01.21267113

**Authors:** Phyllis Awor, Joseph Kimera, Proscovia Athieno, Gloria Tumukunde, Jean Okitawutshu, Antoinette Tshefu, Elizabeth Omoluabi, Aita Signorell, Nina Brunner, Jean-Claude Kalenga, Babatunde Akano, Kazeem Ayodeji, Charles Okon, Ocheche Yusuf, Giulia Delvento, Tristan Lee, Christian Burri, Christian Lengeler, Manuel W. Hetzel

## Abstract

**Background:** In children below 6 years with suspected severe malaria who are several hours from facilities providing parenteral treatment, pre-referral rectal artesunate (RAS) is recommended by the World Health Organization to prevent death and disability. A number of African countries are in the process of rolling out quality-assured RAS for pre-referral treatment of severe malaria at community-level. The success of RAS depends, among other factors, on the acceptability of RAS in the communities where it is being rolled-out. Yet to date, there is limited literature on RAS acceptability. This study aimed to determine the acceptability of RAS by health care providers and child caregivers in communities where quality assured RAS was rolled out.

**Methods:** This study was nested within the comprehensive multi-country observational research project Community Access to Rectal Artesunate for Malaria (CARAMAL). The CARAMAL project was implemented in the Democratic Republic of the Congo (DRC), Nigeria, and Uganda between 2018 and 2020. Data from three different sources were analysed to understand RAS acceptability: Interviews with health workers during three healthcare provider surveys, with caregivers of children under 5 years of age during three household surveys, and with caregivers of children who were recently treated with RAS and enrolled in the CARAMAL Patient Surveillance System.

**Results:** RAS acceptability was high among all interviewed stakeholders in the three countries. After the roll-out of RAS, 97-100% heath care providers in DRC considered RAS medication as very good or good, as well as 98-100% in Nigeria and 93-100% in Uganda. Majority of caregivers whose children had received rectal artesunate for pre-referral management of severe malaria indicated that they would want to get the medication again, if their child had the same illness (99.8% of caregivers in DRC, 100% in Nigeria and 99.9% in Uganda). Further, using data from three household surveys, 67-80% of caregivers whose children had not received RAS considered the medication as useful.

**Conclusion:** RAS was well accepted by health workers and child caregivers in DRC, Nigeria and Uganda. Acceptability is unlikely to be an obstacle to the large-scale roll-out of RAS in the studied settings.

## Introduction

Malaria is one of the leading causes of illness, death, and lost economic productivity globally. It still results in over 400,000 deaths each year, most of which are in children under 5 years of age in sub-Saharan Africa [1]. Most malaria deaths occur in remote settings where there is poor access to formal health facilities and patients do not receive the required treatment and care in a timely manner [1]. Delays in obtaining treatment, particularly for severe disease, can quickly result in lasting sequelae or death. In situations in which parenteral treatment of severe malaria is not available, rectal artesunate (RAS) can be provided as pre-referral treatment for young children. RAS rapidly (within 24 hours) clears 90% or more of malaria parasites, and in a clinical trial it was shown to significantly reduce the risk of death or permanent disability in children less than 6 years of age, who cannot reach a health facility within six hours [2]. The World Health Organization (WHO) therefore recommends treating children less than 6 years of age initially with a single rectal dose of 10 mg artesunate per kilogram of body weight, after which the child should be referred immediately to an appropriate health facility where the full treatment and care package can be provided [3].

RAS is currently available from two WHO pre-qualified suppliers, and it is registered in many malaria-endemic countries, which are at some stage of using rectal artesunate as pre referral treatment for severe malaria [3]. The successful large-scale roll-out of RAS depends, among other factors, on the product’s acceptability in the communities in which it is made available. A number of studies have found the rectal dosage form to be well accepted by parents of sick children in different cultural settings [4]. However, compliance and acceptability of rectal administration may depend on different factors specific to the local context [5, 6]. This study aimed to provide evidence of the acceptability of RAS in communities in the Democratic Republic of the Congo (DRC), Nigeria and Uganda in which RAS has been introduced on a large scale.

## METHODS

### Study design

This study was conducted in the frame of the Community Access to Rectal Artesunate for Malaria (CARAMAL) project, a multi-country operational research aligned with the roll-out of quality assured RAS through established community-based health care providers in DRC, Nigeria and Uganda. In each of the countries, a continuous patient surveillance system (PSS) was set up and Health Care Provider Surveys (HCPS) and Household Surveys (HHS) were conducted annually from 2018 - 2020. More information on the project and research design can be found elsewhere (Lengeler, Burri *et al*., CARAMAL project overview paper, forthcoming).

### Study setting

The study was implemented in the following study areas: Three Health Zones in DRC (Kenge, Kingandu and Ipamu); Three Districts in Uganda (Apac, Kole and Oyam), and three Local Government Areas (Fufore, Mayo-Belwa and Song LGAs) in Adamawa state in Nigeria. The three project countries together share an estimated 42% of total global malaria cases and 39% of total global deaths. In the study areas, health care services are provided by community health workers (CHW) implementing integrated community case management (iCCM) programs, primary health centres (PHC) and referral facilities with inpatient facilities.

### Study populations and data collection

#### Patient Surveillance System

The PSS included children below 5 years of age seeking care for a severe febrile illness episode at the level of community-based care providers (CHW and PHC). Children enrolled in the PSS were followed up at their home 28 days after the initial visit to a CHW or PHC.

#### Health Care Provider Survey

The HCPS included a sample of health workers from all provider levels, including CHW, PHC and referral facilities collected through annual surveys 2018, 2019 and 2020. The selection of the sample was meant to provide a good representation of all types of health workers who treat children with severe febrile illness in the study areas. This analysis was limited to data from CHW and PHC where RAS is administered.

#### Household Survey

For the HHS a stratified 3-stage cluster sampling strategy was utilized. Independent samples were selected annually. The stratification was by Health Zone, District or Local Government Area for each of the three study countries, respectively, and the stages of cluster sampling included the parish level, village level and household level. A sampling interval was used to systematically include surveyed households. In each selected household, the household head and caregivers of children below 5 years of age were eligible to participate in the face-to-face structured interview. The person to be interviewed had to be an adult and able to answer questions about the household characteristics and/or health care of the children below 5 years in that particular household (i.e. usually the primary caregiver).

### Data collection

The surveys were conducted at: baseline (that is prior to introduction of RAS), at midline (about 1 year after introduction of RAS) and at end line (about 2 years after the introduction of RAS). Data was collected by face to face interviews. The interviews were conducted by teams of field interviewers who had received extensive training prior to commencing the survey. Quantitative data collected by field research teams was directly captured on internet/Wi-Fi capable tablets using Open Data Kit (ODK) electronic data collection software.

In the PSS, data was collected through face-to-face interviews with the caregiver of a child that had been enrolled, treated for malaria and followed up 28 days later. For the HHS household heads and caregivers of children under-5 in the household were interviewed.

At each health facility, the field research team conducted an information session with the officer-in-charge upon arrival and, following this, with the health facility staff. The selected health care providers that had consented were then interviewed.

### Analysis

Results from the three data sources are provided, in order to give a holistic view of RAS acceptability among different stakeholders – the healthcare providers (using Healthcare Provider surveys,); caregivers of young children (using the Household Surveys,) as well as caregivers who had direct experience of their child being treated with RAS (using the Patient Surveillance System, PSS). Basic knowledge, attitudes and practices related to RAS, among caregivers and service providers, are presented in this paper. Descriptive analyses of categorical outcomes were performed. For comparisons between years and background characteristics, chi-square statistics are used. Variables related to RAS perception were categorized and interpreted as follows: very good and good = “positive”, neutral, bad and very bad = “neutral/negative”.

### Ethics

The CARAMAL study protocol was approved by the Research Ethics Review Committee of the World Health Organization (WHO ERC, No. ERC.0003008), the Ethics Committee of the University of Kinshasa School of Public Health (No. 012/2018), the Health Research Ethics Committee of the Adamawa State Ministry of Health (S/MoH/1131/I), the National Health Research Ethics Committee of Nigeria (NHREC/01/01/2007-05/05/2018), the Higher Degrees, Research and Ethics Committee of the Makerere University School of Public Health (No. 548), the Uganda National Council for Science and Technology (UNCST, No. SS 4534), and the Scientific and Ethical Review Committee of CHAI (No. 112, 21 Nov 2017). Prior to any provider visit, the relevant local health authorities were informed. All interviews were conducted only after individual written informed consent.

## RESULTS

### Characteristics of study participants – Table 1

#### Household Survey

Across all household surveys, 9332 caregivers of children <5 ears were interviewed. The average age of the caregivers in all the 3 countries was approximately 30 years and the majority of the caregivers were female. See table 1a for more information on the household survey participants. See table 1a for more information on the survey participants.

**Tables:** Rectal Artesunate Acceptability in DRC, Nigeria and Uganda.

House Hold Survey
|  | DRC |  |  | Nigeria |  |  | Uganda |  |  |
| --- | --- | --- | --- | --- | --- | --- | --- | --- | --- |
|  | PRE-RAS<br>2018<br>(N=926) | POST RAS<br>2019<br>(N=947) | POST RAS<br>2020<br>(N=597) | PRE-RAS<br>2018<br>(N=1,018) | POST RAS<br>2019<br>(N=1,386) | POST RAS<br>2020<br>(N=1,399) | PRE-RAS<br>2018<br>(N=1,019) | POST<br>RAS 2019<br>(N=1,020) | POST<br>RAS 2020<br>(N=1,020) |
| Age of caregiver<br>(mean, SD) | 34.5 (13.0) | 30.5 (11.0) | 30.5 (9.3) | 30.1(14.8) | 29.9(14.6) | 30.1(15.34) | 30.9 (10.9) | 31.2<br>(11.6) | 30.9<br>(11.6) |
| Sex of Care giver<br>(Female) | n/a | 89.6 | 76.8 | n/a | 54.7 | 54.1 | 85.7 | 80.3 | 77.1 |

Health care provider survey - Community Health Worker characteristics
|  | n=28 | n=42 | n=31 | n=40 | n=40 | n=40 | n=40 | n=40 | n=40 |
| --- | --- | --- | --- | --- | --- | --- | --- | --- | --- |
| Age (mean, SD) | 45.0 (10.6) | 45.3 (10.4) | 46.8 (9.2) | 31.8 (7.0) | 32.9 (8.5) | 33.4 (7.7) | 40.8 (10.7) | 40.5 (8.9) | 42.6 (9.7) |
| Sex (Female) | 10.7 | 4.8 | 9.7 | 22.5 | 52.5 | 52.5 | 32.5 | 25.0 | 47.9 |
| Number of years<br>as health worker:<br>*mean (Std. Dev.)* | 7.8 (5.1) | 8.3 (6.7) | 7.8 (5.6) | 2.9 (1.1) | 3.9 (1.7) | 4.5 (1.4) | 10.0 (6.2) | 11.1 (5.0) | 11.1 (5.5) |

Health care provider survey - Primary Health Facility (PHC) staff characteristics
|  | n=65 | n=71 | n=37 | n=37 | n=55 | n=49 | n=41 | n=59 | n=53 |
| --- | --- | --- | --- | --- | --- | --- | --- | --- | --- |
| Age (mean, SD) | 44.0 (10.5) | 44.1 (10.9) | 43.9 (9.4) | 40.2 (6.7) | 42.8 (10.2) | 39.9 (10.5) | 34.8 (8.0) | 36.9 (8.2) | 36.9 (8.2) |
| Sex (Female) | 21.4 | 21.1 | 26.9 | n/a | 32.1 | 39.0 | 61.8 | 53.1 | 53.1 |
| Number of years<br>as health worker:<br>*mean (Std. Dev.)* | 14.1 (9.1) | 12.6 (15.1) | 15.0 (9.9) | 11.8 (6.7) | 16.7 (10.7) | 12.2 (9.4) | 9.1 (6.0) | 11.6 (7.6) | 11.6 (7.6) |

Day 28 follow-up of severe malaria patients (2019-2020)
|  | DRC | Nigeria | Uganda |
| --- | --- | --- | --- |
| Number of caregivers whose child received RAS | 1,721 | 144 | 1,780 |
| Age of caregiver (mean, SD) | 31.2 (8.0) | 34.7 (8.9) | 29.3 (9.5) |
| Sex of Care giver (Female) | 38.9 | 73.6 | 84.8 |
| **Highest level of school completed** |  |  |  |
| None | 3.7 | 0.0 | 10.7 |
| Preschool | 3.9 | 0.0 | 41.3 |
| Primary | 23.0 | 31.2 | 37.2 |
| Secondary | 53.4 | 36.1 | 6.3 |
| Some College/University | 14.2 | 3.3 | 1.6 |
| Vocational/technical school | 1.8 | n/a | 2.9 |
| Quranic | n/a | 29.4 | n/a |

#### Health Care Provider Survey

Over the 3 survey rounds, 467 health workers at the primary health care level were interviewed from all the countries. These health workers were about 40 years of age and they had worked for at least 10 years. About 40 community health workers were interviewed annually in each country, and these had been CHWs for over 3 years (range 3-12 years) – table 1a.

#### Patient Surveillance System

Of all the children enrolled in the patient surveillance system during the study period, over 1,700 received RAS as pre referral treatment in DRC and Uganda and 144 in Nigeria – table 1b. The caregivers of these children were mostly female at 73.6% and 84.8% for Nigeria and Uganda, respectively, while those of DRC were mostly male (61.2%). They were mostly aged between 29-35 years. Most caregivers had attended some schooling, up to the primary or secondary school level, in all the three countries.

### RAS acceptability by caregivers of children <5 years – Table 2

During the house hold surveys it was found that knowledge about RAS was very minimal in the communities for the three countries at baseline and gradually improved during the study period rising from 11.1% to 30.9% then to 62.1% for DRC, from 1% to 7.2% then to 13.2% for Nigeria and from 6.3% to 17% and eventually to 47.0% for Uganda. The proportion of children that had received RAS in the communities was low at baseline and slightly increased at endline in the study countries (to 7.8% in DRC, 1.2% in Nigeria and 11.2% in Uganda). A small proportion of caregivers had concerns about the use of RAS – table 3. Some of the concerns related to the use of RAS that were raised included: possibility of side effects as the medicine was thought to be new, unavailability of the medicine and discomfort for the child, due to the rectal route of administration.

**Table 2:** Acceptability of RAS over 3 Household survey rounds: % (95%CI)

|  | DRC |  |  | Nigeria |  |  | Uganda |  |  |
| --- | --- | --- | --- | --- | --- | --- | --- | --- | --- |
|  | PRE-RAS<br>2018<br>(N=926) | POST RAS<br>2019<br>(N=947) | POST RAS<br>2020<br>(N=597) | PRE-RAS<br>2018<br>(N=1,018) | POST RAS<br>2019<br>(N=1,386) | POST RAS<br>2020<br>(N=1,399) | PRE-RAS<br>2018<br>(N=1,019) | POST RAS<br>2019<br>(N=1,020) | POST RAS<br>2020<br>(N=1,020) |
| **Caregiver has heard of RAS before** |  |  |  |  |  |  |  |  |  |
| Yes | 11.1<br>(9.2,13.3) | 30.9<br>(27.9,33.9) | 62.1<br>(58.1,66.1) | 1.4<br>(0.7,2.6) | 8.4<br>(6.3,11.0) | 15.8<br>(13.2,18.8) | 6.3<br>(4.6,8.5) | 16.9<br>(13.1,21.6) | 47.0<br>(42.3,51.7) |
| **Caregiver's child (< 5 years) has received RAS before** |  |  |  |  |  |  |  |  |  |
| Yes | 0.2<br>(0.0,0.8) | 1.8<br>(1.0,2.9) | 4.8<br>(3.2,6.9) | 0.0<br>(0.0,0.4) | 0.8<br>(0.4,1.4) | 1.2<br>(0.7,2.0) | 0.9<br>(0.5,1.7) | 3.1<br>(2.2,4.3) | 5.3<br>(3.2,8.6) |
| **Caregiver has concerns giving rectal artesunate to their child** |  |  |  |  |  |  |  |  |  |
| Yes | 13.7<br>(11.6,16.1) | 11.7<br>(9.7,13.9) | 5.4<br>(3.7,7.5) | n/a | 2.5<br>(1.7,3.6) | 2.4<br>(1.6,3.6) | 3.5<br>(2.3,5.3) | 1.3<br>(0.7,2.6) | 1.1<br>(0.5,2.3) |
| Don't know | 18.4<br>(15.9,21.0) | 28.1<br>(25.3,31.1) | 11.2<br>(8.8,14.1) | n/a | 17.1<br>(14.8,19.7) | 3.2<br>(2.3,4.5) | 13.3<br>(10.6,16.4) | 8.8<br>(6.6,11.7) | 8.5<br>(6.7,10.7) |
| **Caregiver thinks RAS is useful** |  |  |  |  |  |  |  |  |  |
| Yes | 78.2<br>(75.4,80.8) | 67.4<br>(64.3,70.4) | 77.3<br>(73.8,80.7) | n/a | 77.9<br>(75.1,80.4) | 80.1<br>(77.4,82.5) | 72.4<br>(67.3,76.9) | 79.1<br>(75.1,82.6) | 87.5<br>(84.8,89.8) |
| Don't know | 17.8<br>(15.4,20.4) | 28.1<br>(25.3,31.1) | 16.8<br>(13.9,20.0) | n/a | 20.7<br>(18.2,23.4) | 15.2<br>(13.1,17.6) | 23.4<br>(19.1,28.4) | 11.5<br>(9.6,13.8) | 9.5<br>(7.7,11.7) |

n/a – not available (was not collected)

**Table 3:**
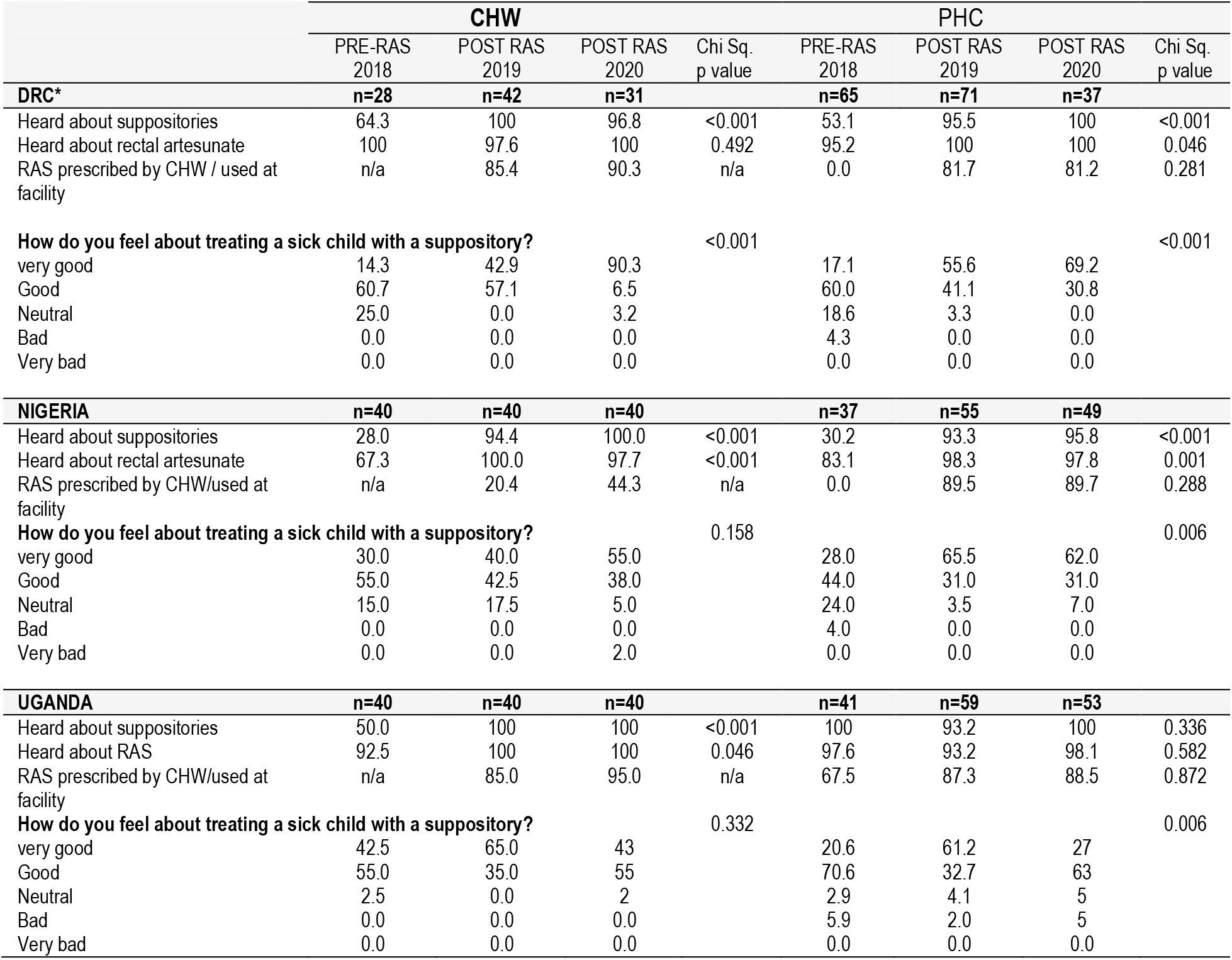
Health care provider Knowledge and Utilization of Rectal Artesunate.

n/a – not available (was not collected)
|  |  |  |  |  |  |  |  |  |
| --- | --- | --- | --- | --- | --- | --- | --- | --- |
|  | **CHW** |  |  |  | **PHC** |  |  |  |
|  | PRE-RAS<br>2018 | POST RAS<br>2019 | POST RAS<br>2020 | Chi Sq.<br>p value | PRE-RAS<br>2018 | POST RAS<br>2019 | POST RAS<br>2020 | Chi Sq.<br>p value |
| **DRC\*** | **n=28** | **n=42** | **n=31** |  | **n=65** | **n=71** | **n=37** |  |
| Heard about suppositories | 64.3 | 100 | 96.8 | <0.001 | 53.1 | 95.5 | 100 | <0.001 |
| Heard about rectal artesunate | 100 | 97.6 | 100 | 0.492 | 95.2 | 100 | 100 | 0.046 |
| RAS prescribed by CHW / used at facility | n/a | 85.4 | 90.3 | n/a | 0.0 | 81.7 | 81.2 | 0.281 |
| **How do you feel about treating a sick child with a suppository?** |  |  |  | <0.001 |  |  |  | <0.001 |
| very good | 14.3 | 42.9 | 90.3 |  | 17.1 | 55.6 | 69.2 |  |
| Good | 60.7 | 57.1 | 6.5 |  | 60.0 | 41.1 | 30.8 |  |
| Neutral | 25.0 | 0.0 | 3.2 |  | 18.6 | 3.3 | 0.0 |  |
| Bad | 0.0 | 0.0 | 0.0 |  | 4.3 | 0.0 | 0.0 |  |
| Very bad | 0.0 | 0.0 | 0.0 |  | 0.0 | 0.0 | 0.0 |  |
| **NIGERIA** | **n=40** | **n=40** | **n=40** |  | **n=37** | **n=55** | **n=49** |  |
| Heard about suppositories | 28.0 | 94.4 | 100.0 | <0.001 | 30.2 | 93.3 | 95.8 | <0.001 |
| Heard about rectal artesunate | 67.3 | 100.0 | 97.7 | <0.001 | 83.1 | 98.3 | 97.8 | 0.001 |
| RAS prescribed by CHW/used at facility | n/a | 20.4 | 44.3 | n/a | 0.0 | 89.5 | 89.7 | 0.288 |
| **How do you feel about treating a sick child with a suppository?** |  |  |  | 0.158 |  |  |  | 0.006 |
| very good | 30.0 | 40.0 | 55.0 |  | 28.0 | 65.5 | 62.0 |  |
| Good | 55.0 | 42.5 | 38.0 |  | 44.0 | 31.0 | 31.0 |  |
| Neutral | 15.0 | 17.5 | 5.0 |  | 24.0 | 3.5 | 7.0 |  |
| Bad | 0.0 | 0.0 | 0.0 |  | 4.0 | 0.0 | 0.0 |  |
| Very bad | 0.0 | 0.0 | 2.0 |  | 0.0 | 0.0 | 0.0 |  |
| **UGANDA** | **n=40** | **n=40** | **n=40** |  | **n=41** | **n=59** | **n=53** |  |
| Heard about suppositories | 50.0 | 100 | 100 | <0.001 | 100 | 93.2 | 100 | 0.336 |
| Heard about RAS | 92.5 | 100 | 100 | 0.046 | 97.6 | 93.2 | 98.1 | 0.582 |
| RAS prescribed by CHW/used at facility | n/a | 85.0 | 95.0 | n/a | 67.5 | 87.3 | 88.5 | 0.872 |
| **How do you feel about treating a sick child with a suppository?** |  |  |  | 0.332 |  |  |  | 0.006 |
| very good | 42.5 | 65.0 | 43 |  | 20.6 | 61.2 | 27 |  |
| Good | 55.0 | 35.0 | 55 |  | 70.6 | 32.7 | 63 |  |
| Neutral | 2.5 | 0.0 | 2 |  | 2.9 | 4.1 | 5 |  |
| Bad | 0.0 | 0.0 | 0.0 |  | 5.9 | 2.0 | 5 |  |
| Very bad | 0.0 | 0.0 | 0.0 |  | 0.0 | 0.0 | 0.0 |  |

Finally, positive perceptions about RAS remained high across the study period, in all the 3 countries (67%-78% in DRC, 78-80% in Nigeria and 72-88% in Uganda.

### RAS acceptability by health workers – Table 3

In 2020, after RAS had been rolled out in the study areas, almost all health providers interviewed had heard about suppositories and rectal artesunate - that is 96.8% in DRC, 100% in Nigeria and 100% in Uganda among community health workers; and 100% in DRC, 95.8% in Nigeria and 100.0% in Uganda for the primary health facilities workers. Most community health workers in DRC and Uganda (90% and 95%) had dispensed RAS and 44% of community health workers in Nigeria had done so by the time of the survey. Over 80% of primary health care providers in all study countries (that is 81% in DRC, 90% in Nigeria and 89% in Uganda) had dispensed RAS. Most health workers interviewed had positive perceptions of RAS, considering it as either very good or good.

Among CHWs and PHCs in DRC, the positive perception of RAS increased from the pre-RAS period of 75% (good or very good) to 96% (good or very good) and 77% (good or very good) to 100% (good or very good) amongst CHWs and PHC providers, respectively. In Nigeria, the increasingly positive perception of RAS among CHWs and PHC was also observed. Whereas health workers in DRC were familiar with RAS even at the pre-RAS survey point (CHW: 100%, PHC: 95.2%), awareness of RAS increased significantly in Nigeria (CHW: from 67.3% to 97.7%, PHC: from 83.1% to 97.8).

In Uganda, a majority of health workers had already heard about suppositories and RAS in particular, prior to the start of this project. After the roll out of RAS, almost all the health workers reported knowledge of RAS.

### RAS acceptability by caregivers of children that recently received RAS for pre-referral management of severe malaria-results from the Patient Surveillance survey – Table 4

The majority of the caregivers (96.6% in DRC, 97.9% in Nigeria and 85.9% in Uganda) were those whose child had received rectal artesunate for the first time.

**Table 4:**
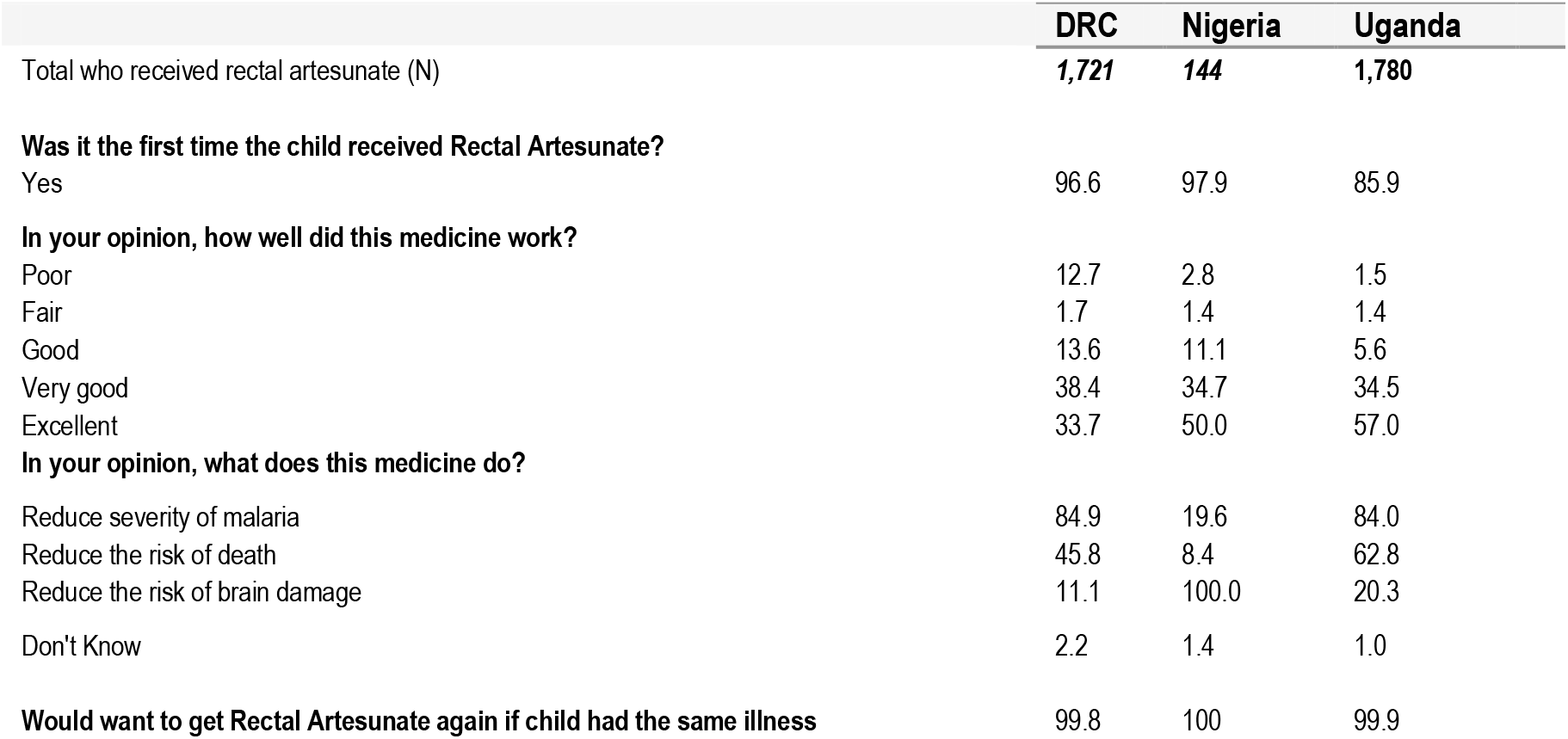
Acceptability of Rectal artesunate by caregivers of children < 5 years with suspected severe malaria Source - Patient surveillance system.

|  | **DRC** | **Nigeria** | **Uganda** |
| --- | --- | --- | --- |
| Total who received rectal artesunate (N) | **1,721** | **144** | **1,780** |
| **Was it the first time the child received Rectal Artesunate?** |  |  |  |
| Yes | 96.6 | 97.9 | 85.9 |
| **In your opinion, how well did this medicine work?** |  |  |  |
| Poor | 12.7 | 2.8 | 1.5 |
| Fair | 1.7 | 1.4 | 1.4 |
| Good | 13.6 | 11.1 | 5.6 |
| Very good | 38.4 | 34.7 | 34.5 |
| Excellent | 33.7 | 50.0 | 57.0 |
| **In your opinion, what does this medicine do?** |  |  |  |
| Reduce severity of malaria | 84.9 | 19.6 | 84.0 |
| Reduce the risk of death | 45.8 | 8.4 | 62.8 |
| Reduce the risk of brain damage | 11.1 | 100.0 | 20.3 |
| Don't Know | 2.2 | 1.4 | 1.0 |
| **Would want to get Rectal Artesunate again if child had the same illness** | **99.8** | **100** | **99.9** |

Almost all caregivers (99.8% in DRC, 100% in Nigeria and 99.9% in Uganda) of the sick children would desire their child to get RAS again if the child had the same illness. When asked about their opinion of how well RAS worked, in DRC, 33. 7% of caregivers rated it as excellent, 38.4% as very good and 13.6% as good. In Nigeria, 50.0% rated RAS as excellent, 34.7% as very good and 11.1% as good. Similarly, in Uganda 57.1% rated the medicine as excellent, 34.5% rated it very good and 5.6% as good while -see Table 4 for more information.

The caregivers were also asked about their opinion regarding what the medicine does. Most of them reported that the medicine is used to reduce severity of malaria that is 84.9% in DRC and 84.0% in Uganda. In Nigeria, the most frequently mentioned answer was that RAS is used to reduce the risk of brain damage (100%).

## DISCUSSION

RAS acceptability was generally high among both community health workers and health workers at primary health care facilities. Two years after the introduction of RAS, 97-100% of health workers in DRC, 98-100% in Nigeria and 93-100% in Uganda considered RAS to be very good or good. These positive perceptions increased over time and with increasing experience of health workers in providing RAS, negative perceptions of RAS remained rare in all three study countries. The high acceptability may be attributed to the fact that there was some use of non-quality assured rectal artesunate within the lower level health facilities, though this was particularly the case in Uganda. Regardless, the study finding of very high proportions of healthcare providers accepting rectal artesunate and rating it as good or very good is encouraging.

RAS use in the communities, as assessed in the household surveys, was initially very low, and only minimally increased during the study period. Despite the low use of RAS, all caregivers interviewed (both those whose children had had an opportunity to receive RAS and those who had only heard about RAS) had very positive perception of RAS as a pre-referral treatment for severe malaria. The caregivers considered RAS to be very useful with many of them stating that RAS was being used to reduce severity of illness and prevent death among the children. Most of the caregivers whose children had previously received RAS were willing to have their child receive RAS if they had the same illness again. Parents whose children had never received RAS also had the correct knowledge and were positive about RAS use.

A few recent studies have also shown high acceptability of RAS [4, 7]. These include one study on adherence to referral advice after use of rectal artesunate for treating children in DRC, where RAS acceptability was found to be 79%, 90% and 98% amongst mothers, community health workers and nurses, respectively [8]. Another study from Uganda in 2016 focused on high compliance with referral advice, after treatment of children with pre-referral rectal artesunate [9].

Acceptability of RAS for pre-referral treatment of malaria was documented to be slightly lower, at 71%, by Hinton *et al*. in their study in 2007 that assessed rectal artesunate acceptability among caregivers in Papua New Guinea [5]. A study by Inthavilay *et al*. in Laos indicated that there were concerns amongst about 40% of the caregivers, related to the rectal route of administration [10]. Our study did not find any strong concerns related to the rectal route of administration.

The strengths of the results presented in this paper are firstly the use of three complementary data sources, namely health care provider surveys, household surveys and the interviews with caregivers of children with suspected severe malaria, many of whom had experience with their children receiving RAS. The results from these data sources are consistent, showing high acceptability of rectal artesunate. Secondly, we utilize large data sets, with thousands of patients and caregivers included. Thirdly, we include data from three countries with different cultural and health systems environments but a very high malaria burden. The consistent information obtained across the data sources outweighs any possible recall bias.

In conclusion, the results from our studies confirm high acceptability of using rectal artesunate in children with signs of severe malaria in DRC, Nigeria and Uganda. Acceptability is therefore unlikely to be an obstacle to the large-scale roll-out of RAS in the studied settings. Together with previous evidence, the rectal formulation does not appear to be a concern to health workers of caregivers of sick children in a variety of cultural settings.

## Data Availability

All data produced in the present study are available upon reasonable request to the authors

## Notes

### Competing Interest Statement

The authors have declared no competing interest.

### Funding Statement

This study was funded by Unitaid

### Author Declarations

The CARAMAL study protocol was approved by the Research Ethics Review Committee of the World Health Organization (WHO ERC, No. ERC.0003008), the Ethics Committee of the University of Kinshasa School of Public Health (No. 012/2018), the Health Research Ethics Committee of the Adamawa State Ministry of Health (S/MoH/1131/I), the National Health Research Ethics Committee of Nigeria (NHREC/01/01/2007-05/05/2018), the Higher Degrees, Research and Ethics Committee of the Makerere University School of Public Health (No. 548), the Uganda National Council for Science and Technology (UNCST, No. SS 4534), and the Scientific and Ethical Review Committee of CHAI (No. 112, 21 Nov 2017).

